# The prevalence and patterns of illicit alcohol consumption in twelve African countries: Evidence from WHO STEPS Surveys (2014–2019)

**DOI:** 10.64898/2026.01.16.26344252

**Authors:** Cosmas Zyambo, Kennedy Niwagaba, Wingston Felix Ng’ambi

## Abstract

**Introduction:** Unrecorded alcohol, untaxed and often illicitly produced, poses major public health risks in Africa due to its unregulated nature, affordability, and widespread availability among vulnerable populations. Cultural practices, poverty, and weak policy enforcement contribute to its high use, especially among young people and rural communities.

**Methods:** This study used secondary data from the WHO STEPS conducted between 2014 and 2019 across twelve African countries: Algeria, Benin, Botswana, Eswatini, Ethiopia, Kenya, Malawi, Morocco, São Tomé and Príncipe, Sudan, Uganda, and Zambia. Multivariable logistic regression examined demographic and socioeconomic factors associated with illicit alcohol consumption, while quantile regression assessed variations in ethanol intake. Analyses were adjusted for complex survey design to ensure nationally representative and comparable estimates across countries.

**Results:** The overall prevalence was 8.3%, ranging from 0.1% in Morocco to 12.9% in Zambia. Men were more likely to consume illicit alcohol than women, with higher use observed among older adults, rural residents, and individuals with lower education levels. The median ethanol intake among illicit alcohol users was 62.5 g (IQR: 31.2–156.2 g), indicating heavy consumption among certain groups. Multivariable analysis confirmed significant associations between illicit alcohol use and socioeconomic disadvantage, highlighting deep inequalities that shape alcohol access and consumption patterns across the region.

**Conclusion:** The study highlights the urgent need for stronger surveillance, enforcement, and community-based prevention strategies. Policymakers should integrate alcohol control into broader health and social protection programs, ensuring safer and more affordable alternatives. Future work should include longitudinal monitoring, biochemical validation, and cross-border collaboration to address unrecorded alcohol trade and reduce its impact across the region.

## INTRODUCTION

Globally, alcoholic beverages are categorized as either recorded or unrecorded (also referred to as illicit). Recorded alcohol is legally produced, regulated, monitored, and sold through official channels. Unrecorded or illicit alcohol, by contrast, encompasses a range of products including: 1) Alcohol produced by unlicensed individuals or industries; 2) Homemade or home-brewed alcohol; 3) Products that bypass regulatory oversight; 4) Alcohol smuggled across national borders;5) Consumption within unregistered or unauthorized jurisdictions; 6) Substances not intended for consumption, such as surrogate alcohols (e.g., hand sanitizer or aftershave) (1-4) ^1,2,3,4^,. This includes homemade brews, illicitly produced alcohol, and products not intended for human consumption but consumed as substitutes (e.g., industrial spirits)^5^. Globally, it is estimated that approximately 25% of all alcohol consumed is Illicit ^6 7^ with Africa’sconsumption of illicit alcohol often exceeding that of recorded alcohol in both frequency and quantity ^8 9^. These beverages are often produced in unregulated environments without adherence to safety standards, increasing the risk of contamination and acute toxicity. Moreover, their affordability and wide availability make them accessible to vulnerable populations, including youth and low-income individuals, exacerbating alcohol-related harms such as liver disease, mental health disorders, and increased risk of injury and violence^10 8 11^.

Alcohol consumption is often shaped by complex social, religious, economic, and cultural dynamics^12^. While Illicit alcohol accounts for approximately 21.9% of total per capita alcohol consumption in Europe, this figure increases significantly to 56.2% in the Eastern Mediterranean and 69% in Southeast Asia^13^. In some countries, Illicit alcohol constitutes a large share of overall consumption-accounting for 59% in Bhutan, 44.4% in Kuwait, 42.3% in Uganda, 40.1% in the Republic of Moldova.^14^ and 71.2% in Zambia ^15^. While overall per capita alcohol use in many African countries may appear lower than in high-income settings^13^, a disproportionate share of this consumption comes from Illicit sources. In some regions, traditional brewing practices and limited enforcement of alcohol policies contribute to high levels of Illicit consumption. Additionally, gender norms and poverty play critical roles in shaping drinking behaviors ^15–17^, with men and rural populations often reporting higher use of Illicit alcohol^15^. The health burden is notably higher among men, with 7.4% of all male deaths linked to alcohol consumption, compared to 1.4% among women^18^. These patterns not only complicate public health surveillance but also undermine efforts to implement effective alcohol taxation, regulation, and harm reduction policies.

The WHO STEPwise approach to Surveillance (STEPS) provides a vital tool for understanding the landscape of Illicit alcohol use across African countries. As a standardized, nationally representative survey focused on non-communicable disease risk factors, STEPS allows for cross-country comparisons and trend analysis over time. By collecting data on alcohol consumption, including types and sources, STEPS enables researchers and policymakers to identify population-level patterns and assess the burden of unrecorded alcohol use. This evidence is essential for designing targeted interventions, informing policy decisions, and ultimately reducing the health and social harms associated with unrecorded alcohol in Africa.

This study aims to examine the prevalence, patterns, and determinants of Illicit alcohol consumption across African countries using data from WHO STEPS surveys conducted between 2014 and 2019, with the goal of informing policy and public health interventions to reduce alcohol-related harm. Specifically, it seeks to estimate the prevalence of Illicit alcohol use and identify key demographic, socioeconomic, and behavioral factors associated with its consumption; compare patterns across countries and regions to uncover geographic disparities and trends; assess temporal changes in countries with multiple rounds of STEPS data; and provide policy-relevant recommendations to support effective regulation and mitigation of the health impacts of unrecorded alcohol in the African context.

## METHODS

### Data source

The WHO STEPwise approach to NCD risk factor surveillance (STEPS) is a standardized methodology developed to collect, analyze, and disseminate data on key non-communicable disease risk factors. Between 2014 and 2019, STEPS surveys were conducted across multiple African countries to monitor trends in behaviors such as tobacco use, physical inactivity, diet, and alcohol consumption. The surveys follow a three-step model: Step 1 collects demographic and behavioral information through structured interviews; Step 2 involves physical measurements (e.g., height, weight, blood pressure); and Step 3 includes biochemical assessments such as blood glucose and cholesterol levels. This structured approach ensures consistency in data collection and facilitates international comparability. During the 2014–2019 period, WHO STEPS surveys were conducted in various African countries namely Algeria, Benin, Botswana, Eswatini, Ethiopia, Kenya, Malawi, Morocco, São Tomé and Príncipe, Sudan, Uganda, and Zambia. This regional spread across North, East, and Southern Africa enriched the analysis by reflecting varying alcohol consumption patterns and health behaviors. These datasets was accessed on the 30^th^ of June 2023 and authors had no access to information that could identify individual participants during or after data collection.

### Variables and measurements

The individual data from the country WHO STEPS were combined into a single dataset for analysis ^19^. The dataset had the outcome and predictor variables as described below.

#### Outcome variable of interest

The main outcome variable was illicit (unrecorded) alcohol consumption participants that responded to the question: “During the past 7 days, did you consume any homebrewed alcohol, any alcohol brought over the border or from another country, any alcohol not intended for drinking, or other untaxed alcohol?”. In WHO STEPS; the participants were asked, “During the past 7 days, did you consume any homebrewed alcohol, any alcohol brought over the border or from another country, any alcohol not intended for drinking (e.g., alcohol-based medicines, perfumes, aftershaves), or other untaxed alcohol?”. Those who responded “Yes” were coded as 1, indicating illicit alcohol consumption, while those who responded “No” were coded as 0, indicating no illicit consumption. Beyond prevalence, we also examined the volume of ethanol in the unrecorded alcohol consumed. To estimate the total volume of pure ethanol consumed from unrecorded alcohol, we applied alcohol-by-volume (ABV) conversion factors to each beverage type reported in the WHO STEPS survey ^20^. Using standardized drink sizes and the density of ethanol (0.789 g/mL), we calculated the grams of ethanol consumed per alcohol type. Specifically, consumption volumes were estimated as follows: 25 mL per spirit drink, 330 mL per beer or wine, 150 mL per imported alcohol, 30 mL for industrial alcohol not intended for drinking, and 50 mL for other untaxed or unspecified alcohol. Each reported drink count was multiplied by its corresponding ABV and ethanol density to derive the total ethanol content per participant. The resulting estimates provided a continuous measure of unrecorded alcohol consumption volume, adjusted for beverage type and concentration, enabling comparison across individuals and subgroups.

#### Explanatory variables

We included key sociodemographic characteristics such as age, sex (male or female), and place of residence (urban or rural). Socioeconomic status was measured using employment type (government employee, non-government employee, self-employed, or unpaid/retired), highest level of education (none, primary, secondary, or tertiary), and marital status (never married, currently married, or formerly married). In addition to demographics, we examined several modifiable behavioural risk factors captured in the WHO STEPS 2024 questionnaire. These included current tobacco use (never, previous or current), reported salt intake (high or low), and fruit and vegetable consumption (low or adequate). Physical activity levels were also assessed and categorised as low or high based on participants’ reported frequency and intensity of activity. The AUDIT-C is scored on a scale of 0–12 (scores of 0 reflect no alcohol use). Alcohol drinking is based on the AUDIT-C tool (AUDIT-C score ≤ 2 for women and ≤ 3 for men was classified as ‘harmful alcohol use’ (no); AUDIT-C score ≥ 3 for women and ≥4 for men was classified as ‘harmful alcohol use’ (yes)^16,21^. Responses were coded as yes or no to indicate the presence of hypertension or diabetes, respectively.

### Statistical Analysis

Data analysis followed a structured, multi-stage approach to capture both the prevalence and volume of unrecorded alcohol consumption. All analyses accounted for the complex sampling design of the WHO STEPS surveys by applying sample weights, primary sampling units, and strata to generate nationally representative estimates. The analysis was conducted using R software (version 3.4.5). We first estimated overall and subgroup-specific weighted prevalences of unrecorded alcohol use. To identify key sociodemographic, behavioural, and clinical correlates, we conducted bivariate analyses followed by multivariable logistic regression ^22^. A backward stepwise approach, guided by the Wald test at a 0.05 significance level, was used to build the final model, reporting adjusted odds ratios (AORs) with 95% confidence intervals.

To complement the prevalence analysis, we assessed consumption volume by estimating the median grams of ethanol consumed from unrecorded sources. We further examined how consumption patterns varied by beverage type (such as spirits, beer, or wine) and across population subgroups using quantile regression at the median (τ = 0.5). This approach provided a robust method for modelling the determinants of consumption volume while minimizing the effect of outliers. Results were visualized through clear, publication-ready graphics, including forest plots to display prevalence estimates and confidence intervals, and dot plots to illustrate variations in median consumption across key factors. A p-value of less than 0.05 was considered statistically significant.

### Ethical approval

All participants in the original WHO STEPS surveys provided written informed consent prior to data collection. For this study, permission to access and use the data for secondary analysis was formally obtained from the World Health Organization, which manages and funds the STEPS surveys. The anonymized datasets were accessed and downloaded from the WHO NCD Microdata Repository (https://extranet.who.int/ncdsmicrodata/index.php/catalog/629). No personal identifiers were included in the dataset, ensuring full protection of participant privacy and confidentiality throughout the analysis.

## RESULTS

### Characteristics of study participants

Geographically, the study covered 12 African countries, offering a broad perspective on unrecorded alcohol consumption across diverse cultural and social settings. Ethiopia contributed the largest share of participants (23.6%), followed by Algeria (21.6%) and Kenya (16.1%) (Table 1). Smaller proportions came from countries such as Benin, Morocco, and São Tomé and Príncipe.

**Table 1.** Participant characteristics from 12 African countries that were included in the study: 2014–2019.

| Characteristics | N | % |
| --- | --- | --- |
| **Total** | **46040** | **100.0** |
| **Age group (years)** |  |  |
| 15–19 | 2655 | 8.0 |
| 20–24 | 6182 | 18.1 |
| 25–29 | 6755 | 18.9 |
| 30–34 | 6664 | 12.7 |
| 35–39 | 6111 | 11.6 |
| 40–44 | 5000 | 10.0 |
| 45–49 | 3966 | 7.0 |
| 50–54 | 3233 | 5.7 |
| 55–59 | 2313 | 3.8 |
| 60–64 | 1733 | 2.2 |
| 65+ | 1428 | 1.9 |
| **Sex** |  |  |
| Female | 27363 | 47.7 |
| Male | 18677 | 52.3 |
| **Residence** |  |  |
| Rural | 25563 | 59.1 |
| Urban | 20477 | 40.9 |
| **Highest education level** |  |  |
| None | 15485 | 28.4 |
| Primary | 15111 | 35.4 |
| Secondary | 11186 | 25.9 |
| Tertiary | 4258 | 10.4 |
| **Occupation** |  |  |
| Government employee | 3677 | 8.6 |
| Non-government employee | 3735 | 9.0 |
| Employed (Informal) | 19599 | 46.4 |
| Not in the Labor Force | 19029 | 36.0 |
| **Marital status** |  |  |
| Never married | 10310 | 25.3 |
| Currently married | 32366 | 69.1 |
| Formerly married | 3364 | 5.5 |
| **Country** |  |  |
| Algeria | 5547 | 21.6 |
| Benin | 4728 | 2.1 |
| Botswana | 1873 | 0.4 |
| Eswatini | 2384 | 0.3 |
| Ethiopia | 7202 | 23.6 |
| Kenya | 4036 | 16.1 |
| Malawi | 3250 | 6.6 |
| Morocco | 4462 | <0.1 |
| São Tomé & Príncipe | 2030 | 0.2 |
| Sudan | 3949 | 12.1 |
| Uganda | 3384 | 10.9 |
| Zambia | 3195 | 6.1 |
| **Diabetes** |  |  |
| No | 44906 | 97.5 |
| Yes | 1134 | 2.5 |
| **Hypertension** |  |  |
| No | 40817 | 92.1 |
| Yes | 5223 | 7.9 |
| **High salt intake** |  |  |
| No | 32342 | 67.1 |
| Yes | 13698 | 32.9 |
| **Low fruit intake** |  |  |
| No | 10816 | 18.8 |
| Yes | 35224 | 81.2 |
| **Low vegetable intake** |  |  |
| No | 24429 | 47.2 |
| Yes | 21611 | 52.8 |
| **Physical activity level** |  |  |
| Low | 8757 | 15.4 |
| Moderate | 9670 | 19.4 |
| High | 27613 | 65.2 |
| **Tobacco use** |  |  |
| Never | 39748 | 83.0 |
| Previous | 2499 | 6.6 |
| Current | 3793 | 10.4 |
| **Harmful alcohol use** |  |  |
| No | 44604 | 96.9 |
| Yes | 1436 | 3.1 |
| **Survey year** |  |  |
| 2014 | 7641 | 11.6 |
| 2015 | 15966 | 41.8 |
| 2016 | 9496 | 33.8 |
| 2017 | 10907 | 12.7 |
| 2019 | 2030 | 0.2 |

The study population represented a wide age range, with the majority being young adults aged 20-34 years. The 25-29 age group accounted for the largest segment (18.9%), closely followed by those aged 20-24 (18.1%), while participants aged 65 years and above made up less than 2% (Table 1). Men constituted a slightly higher proportion (52.3%) than women (47.7%), and most respondents lived in rural areas (59.1%). Marital status showed that 69.1% were currently married, 25.4% had never married, and 5.5% were formerly married. Educational attainment was generally low, with 28.4% having no formal education and 35.4% completing only primary level, while just 10.4% had tertiary education. Employment data revealed that nearly half of the participants were self-employed (46.4%), and over one-third were unpaid workers or retired (36%).

Health and behavioral indicators revealed important trends. As shown in Table 1, the prevalence of diabetes and hypertension was relatively low, at 2.5% and 7.9% respectively, but behavioral risk factors were widespread. More than 80% of participants consumed insufficient fruit, and over half reported low vegetable intake. Most individuals (65.2%) engaged in high physical activity, although 15.4% had low activity levels. Tobacco use was minimal, with 83% reporting they had never smoked, while harmful alcohol use was observed among 3.1% of respondents. The data, primarily drawn from WHO STEPS surveys conducted between 2014 and 2017 - with 2015 as the peak year - provided a robust understanding of health-related behaviors across diverse African populations, highlighting key areas for intervention and policy attention.

### Prevalence of illicit alcohol use in Africa

The overall prevalence of illicit alcohol use across the twelve African countries was 8.3%. The highest rates in Zambia (12.9%), Uganda (9.0%), and São Tomé and Príncipe (8.5%), and minimal use in Morocco (0.1%) and Algeria (0.2%). Malawi reported a relatively low prevalence of just over 1% (see Figure 1). Prevalence increased with age, from 5.5% among adolescents (15–19 years) to 15.7% in adults aged 65 and older, and was higher in males (13.0%) than females (5.5%) (Figure 1). Participants with no formal education had higher prevalence (17.8%) than those with tertiary education (3.4%), and self-employed individuals were more likely to use illicit alcohol (12.9%) than government or non-government employees. Marital status and residence also mattered: formerly married participants had the highest prevalence (13.8%), and rural residents reported higher use (12.6%) than urban dwellers (4.9%).

**Figure 1:**
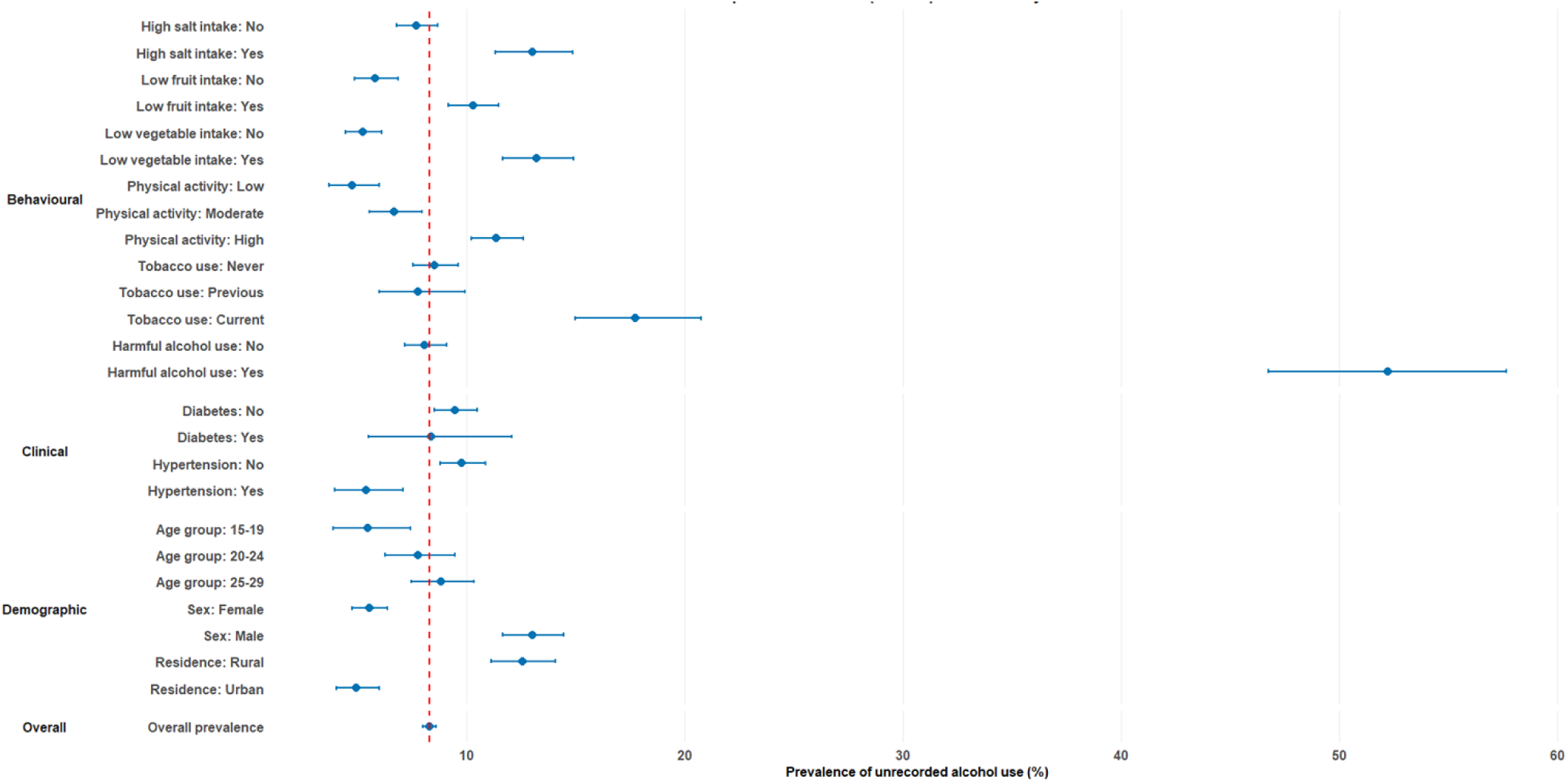
Prevalence of illicit alcohol consumption in twelve African countries that were included in the study: 2014–2019

Behavioral and lifestyle factors were strong correlates. High salt intake and low fruit or vegetable consumption were linked to elevated prevalence (13.0% and 13.2%, respectively). Tobacco use and harmful alcohol consumption were also associated with higher illicit alcohol use, with 17.7% of current smokers and 52.2% of harmful alcohol users reporting consumption. Physical activity showed an unexpected pattern, with higher prevalence among those with high activity (11.4%) than low activity (4.9%). Diabetes and hypertension were not strongly associated with use. Trends over time suggested rising uptake, from 14.8% in 2014 to 22.8% in 2019 (Figure 1), highlighting substantial variation across countries, demographic groups, and behavioral profiles.

### Determinants of illicit alcohol use in Africa

After adjustment for potential confounders, several sociodemographic and behavioural factors were significantly associated with unrecorded alcohol use (Figure 2). The likelihood of consuming unrecorded alcohol increased progressively with age. Compared with individuals aged 15–19 years (reference), those aged 20–24 years had approximately double the odds of illicit alcohol use (AOR = 2.05; 95% CI: 1.16–3.63; p = 0.014). The odds continued to rise with age, peaking among individuals aged 55–59 years (AOR = 4.39; 95% CI: 2.30–8.38; p < 0.001) and remaining elevated in the 65+ group (AOR = 3.61; 95% CI: 1.33–9.83; p = 0.012). Men were nearly three times more likely than women to use unrecorded alcohol (AOR = 2.88; 95% CI: 2.20–3.76; p < 0.001).

**Figure 2:**
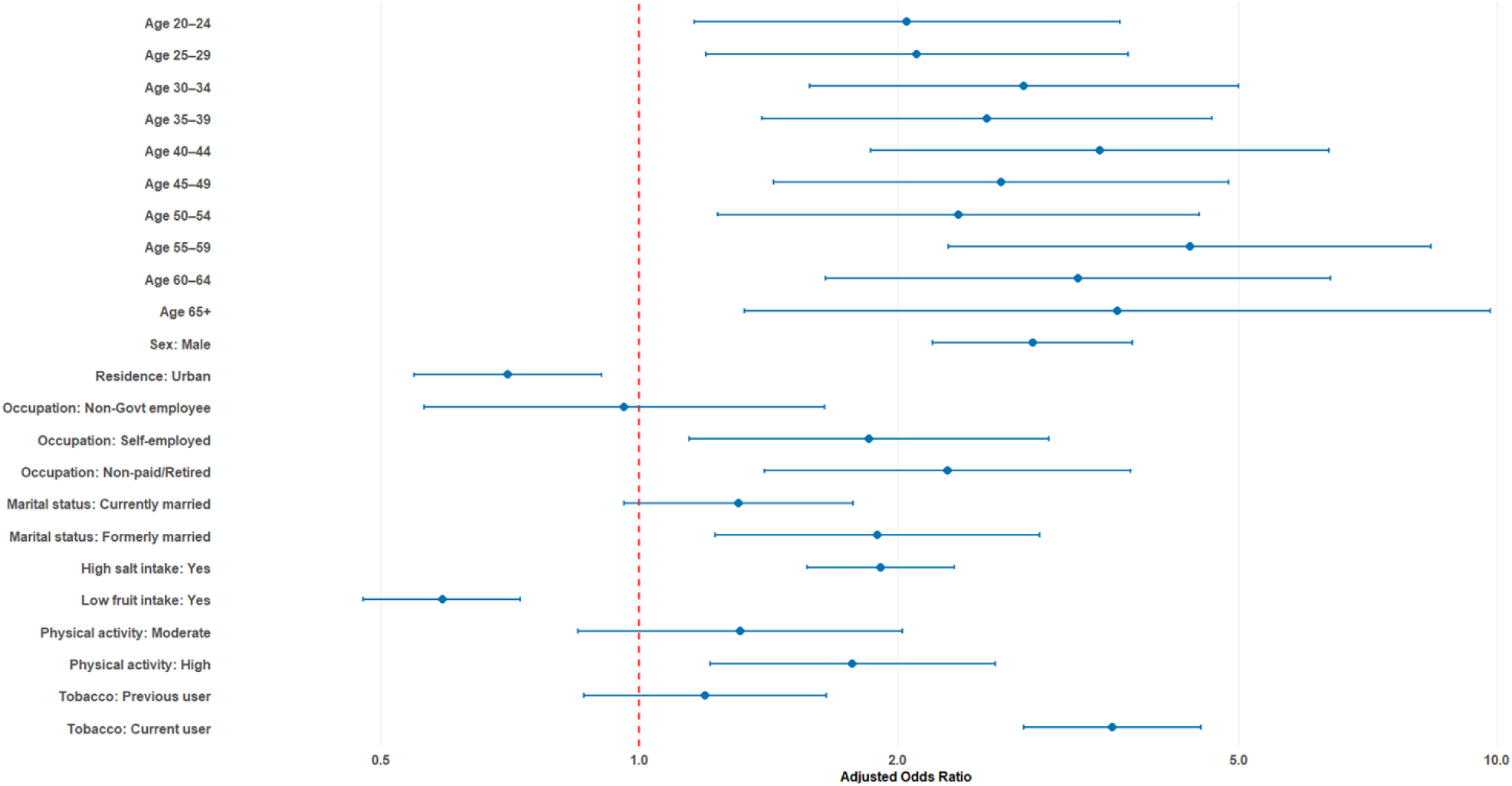
Determinants of illicit alcohol use in twelve African countries that were included in the study: 2014–2019

Residence and marital status also showed significant associations as shown in Figure 2. Individuals living in urban areas had lower odds of unrecorded alcohol use than those in rural areas (AOR = 0.70; 95% CI: 0.55–0.91; p = 0.006). Marital status showed a gradient, with formerly married participants being almost twice as likely as never-married individuals to consume unrecorded alcohol (AOR = 1.90; 95% CI: 1.23–2.93; p = 0.004). Employment status further differentiated risk: self-employed individuals (AOR = 1.85; 95% CI: 1.14–3.01; p = 0.012) and those not in paid work or retired (AOR = 2.29; 95% CI: 1.40–3.74; p = 0.001) exhibited significantly higher odds compared to government employees.

Health-related behaviours were strongly correlated with unrecorded alcohol use (see Figure 2). Participants reporting high salt intake were nearly twice as likely to use unrecorded alcohol (AOR = 1.91; 95% CI: 1.57–2.33; p < 0.001), while those with low fruit intake showed lower odds (AOR = 0.59; 95% CI: 0.48–0.73; p < 0.001). Physical activity displayed a non-linear association: moderate levels were not significantly related (AOR = 1.31; 95% CI: 0.85–2.03; p = 0.222), but high activity levels were linked to greater odds of use (AOR = 1.77; 95% CI: 1.21–2.60; p = 0.003). Smoking status was a particularly strong correlate; current smokers had more than threefold higher odds of unrecorded alcohol use compared to non-smokers (AOR = 3.56; 95% CI: 2.81–4.52; p < 0.001), while former smokers did not differ significantly (AOR = 1.19; 95% CI: 0.86–1.65; p = 0.285).

### Average ethanol consumption from illicit alcohol in Africa

#### Average ethanol consumption from all illicit alcohol

Among consumers of illicit alcohol, intake was high and varied considerably, with a median of 62.5 (IQR: 31.2-156.2) grams of ethanol, indicating a significant subset of heavy drinkers. Consumption patterns revealed clear demographic and behavioral divides. Men consumed roughly five times more than women, and rural residents consumed over 2.5 times more than their urban counterparts. Intake was strongly associated with socioeconomic status, being substantially higher among individuals with no formal education and the self-employed compared to those with tertiary education or formal employment. Furthermore, key lifestyle risk factors; such as current smoking, high salt intake, and low fruit/vegetable consumption; were strongly linked to higher illicit alcohol use. In contrast, clinical conditions like diabetes and hypertension showed no strong association, underscoring that socio-demographic and behavioral factors are the primary drivers of unrecorded alcohol consumption.

#### Average ethanol consumption from illicit spirits

The low consumption of spirits among unrecorded alcohol users, with a median intake of 0.0 g (IQR: 0.0–15.8 g), suggesting that most consumed little or no spirits-derived ethanol (see Figure 3). Among unrecorded alcohol consumers, spirits intake was generally low but varied across age, gender, and lifestyle. Consumption increased with age, from 0.39 g (95% CI: 0.00–0.77) among adolescents 15–19 years to 4.61 g (95% CI: 2.23–7.00) in adults 55–59 years. Men consumed substantially more than women, 3.57 g (95% CI: 3.05–4.09) versus 0.42 g (95% CI: 0.30–0.54), and rural residents reported higher intake than urban counterparts, 2.94 g (95% CI: 2.50–3.38) versus 0.79 g (95% CI: 0.59–0.99). Education and occupation influenced consumption: individuals with no formal education and self-employed participants had the highest intake, while those with tertiary education or government/non-government employment consumed less. Formerly married participants also reported higher spirits consumption than currently married or never-married individuals. Lifestyle factors such as high salt intake, low fruit and vegetable consumption, high physical activity, and smoking were associated with increased spirits intake, with current smokers averaging 8.07 g (95% CI: 6.12–10.03). Clinical conditions like diabetes or hypertension had minimal influence, highlighting socio-demographic and behavioral factors as the main drivers of spirits consumption.

**Figure 3:**
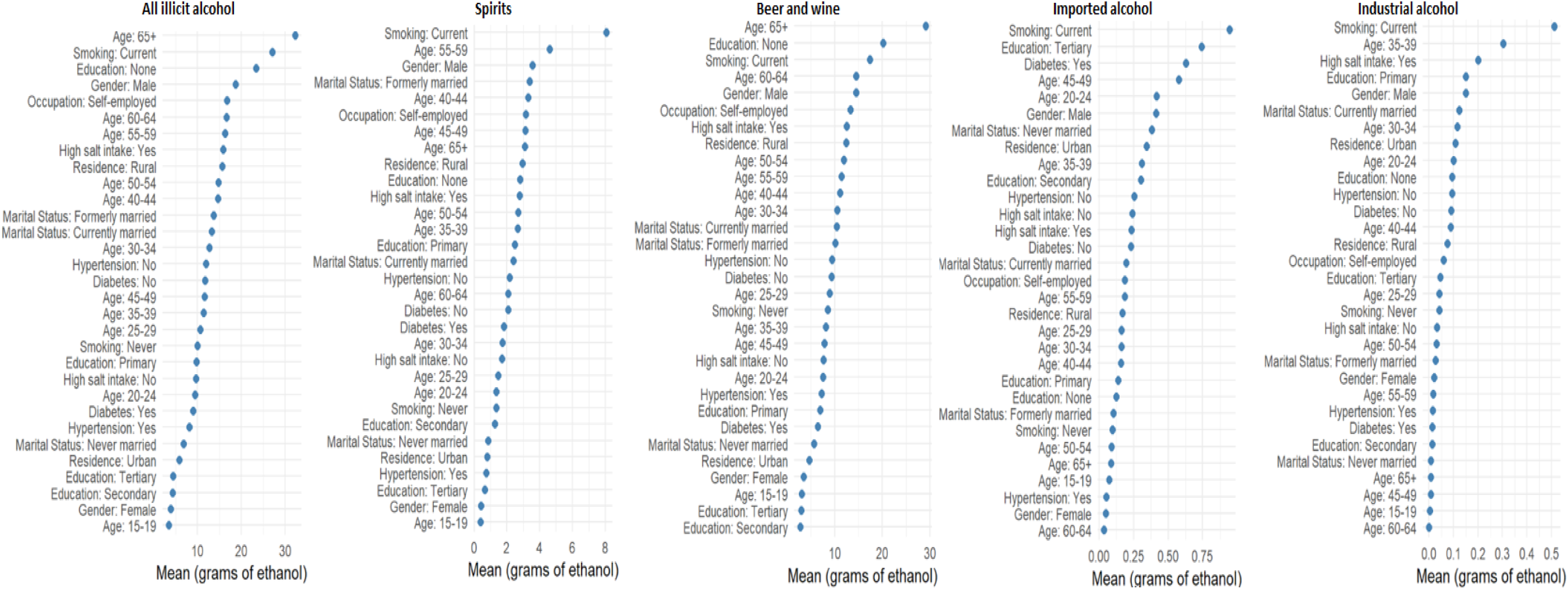
Average ethanol consumption by type of unrecorded alcohol across socio-demographic groups in twelve African countries, 2014–2019

**Figure 4:**
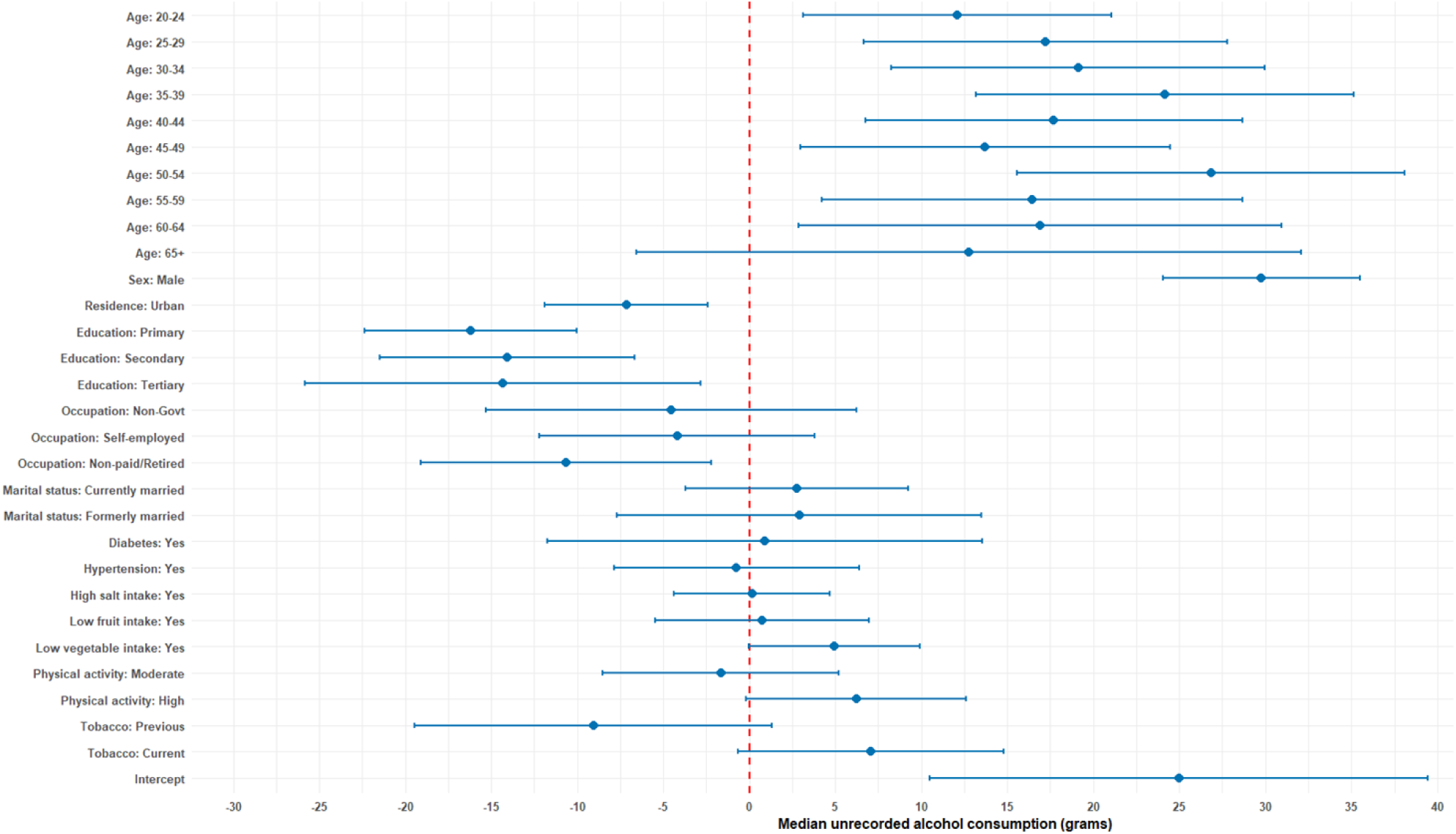
Determinants of median illicit alcohol consumption in twelve African countries that were included in the study: 2014–2019

#### Average ethanol consumption from illicit beer and wine

The ethanol intake from beer and wine among unrecorded alcohol users had a median of 46.9 g (IQR: 15.6–109.4 g), indicating moderate but variable consumption levels across individuals (see Figure 3). Among participants consuming unrecorded beer and wine, ethanol intake showed notable variation across socio-demographic and lifestyle factors. Intake increased with age, from 3.02 g (95% CI: 1.68–4.36) in adolescents 15–19 years to 29.1 g (95% CI: 0.3–57.9) in adults aged 65 years and above. Males consumed considerably more than females, 14.5 g (95% CI: 13.5–15.5) versus 3.45 g (95% CI: 3.0–3.9), while rural residents reported higher intake than urban residents, 12.4 g (95% CI: 11.0–13.8) versus 4.61 g (95% CI: 3.4–5.8). Lower educational attainment and self-employment were associated with higher consumption, whereas participants with tertiary education or government/non-government employment had lower intake. Formerly married participants also showed elevated beer and wine use compared to currently married or never-married individuals. Behavioral factors such as high salt intake, low fruit and vegetable consumption, high physical activity, and smoking correlated with increased intake, with current smokers consuming 17.4 g (95% CI: 10.2–24.6). Clinical conditions such as diabetes or hypertension had minimal impact, suggesting that socio-demographic and lifestyle factors are stronger drivers of beer and wine consumption in this population.

#### Average ethanol consumption from illicit imported Alcohol

The imported alcohol showed the lowest ethanol intake among unrecorded alcohol users, indicating that such beverages contributed negligibly to total unrecorded alcohol consumption (see Figure 3). Ethanol consumption from imported unrecorded alcohol was generally low across the population. Intake was minimal among adolescents aged 15–19 at 0.073 g (95% CI: 0.0–0.18), increasing slightly in adults 20–24 years to 0.415 g (95% CI: 0.0–1.03), and remained below 1 g in most age groups, with a peak of 0.743 g (95% CI: 0.0–1.84) among those with tertiary education. Males consumed substantially more than females, 0.411 g (95% CI: 0.16–0.66) versus 0.049 g (95% CI: 0.02–0.08), and urban residents showed slightly higher intake than rural residents, 0.342 g (95% CI: 0.06–0.62) versus 0.167 g (95% CI: 0.06–0.27). Participants with lower educational attainment or self-employment exhibited modestly higher intake, whereas government or non-government employees had lower consumption. Lifestyle factors such as high salt intake, low fruit and vegetable intake, high physical activity, and current smoking were associated with slightly elevated intake, while clinical conditions including diabetes and hypertension showed minimal influence. Overall, these findings suggest that imported unrecorded alcohol contributes only marginally to total ethanol consumption in the population, with socio-demographic and behavioral factors shaping exposure.

#### Average ethanol consumption from illicit industrial alcohol

The industrial ethanol consumption was also extremely low as shown in Figure 3. Ethanol intake from industrial alcohol was very low across all population groups. Among adolescents aged 15–19, the mean intake was 0.005 g (95% CI: 0.0–0.013), increasing slightly to 0.10 g (95% CI: 0.0–0.22) among those aged 20–24 and peaking at 0.30 g (95% CI: 0.0–0.87) in adults aged 35–39. Older adults consumed negligible amounts, with nearly zero intake beyond age 60. Males recorded higher consumption, averaging 0.15 g (95% CI: 0.01–0.29), compared to females at 0.02 g (95% CI: 0.0–0.04). Urban dwellers consumed slightly more than rural residents, 0.11 g (95% CI: 0.0–0.27) versus 0.07 g (95% CI: 0.03–0.12). Those with primary education and non-government employment showed slightly elevated levels, while the tertiary educated and government workers had minimal exposure. Behavioural risk factors also showed a small gradient, with those reporting high salt intake, high physical activity, and current smoking exhibiting somewhat higher mean intakes, 0.20 g (95% CI: 0.0–0.42), 0.13 g (95% CI: 0.02–0.23), and 0.51 g (95% CI: 0.0–1.17) respectively. These findings suggest that industrial alcohol use is rare and contributes negligibly to total ethanol intake, though isolated consumption among certain subgroups especially working-age males and high-risk behaviour clusters may still warrant attention from public health authorities.

#### Factors influencing average levels of illicit ethanol intake

The quantile regression (τ = 0.5) showed that the median ethanol intake from illicit alcohol varied across age, gender, education, and residence. Consumption increased with age, peaking among adults aged 50–54 years (β = 26.8, 95% CI: 15.7–38.0, p < 0.001). Men consumed significantly more than women (β = 29.8, 95% CI: 24.7–34.9, p < 0.001), while urban residents reported lower median intake (β = –7.1, 95% CI: –11.9 to –2.4, p = 0.003). Education had a protective effect, with primary (β = –16.2, 95% CI: –21.8 to –10.6, p < 0.001), secondary (β = –14.1, 95% CI: –21.5 to –6.6, p < 0.001), and tertiary education (β = –14.3, 95% CI: –24.3 to –4.4, p = 0.005) associated with lower consumption. High physical activity (β = 6.2, 95% CI: 0.4–12.0, p = 0.036) and living in violent settings (β = 5.0, 95% CI: 0.7–9.2, p = 0.022) were linked to greater use. Overall, the median level of illicit ethanol consumption was highest among middle-aged, less-educated rural men.

## DISCUSSION

Illicit alcohol consumption remains a major yet often overlooked public health concern across Africa, contributing significantly to the region’s alcohol-related disease burden and social harm. In this study, the overall prevalence of illicit alcohol use was 8.3%, with notable country differences: ranging from just 0.1% in Morocco to nearly 13% in Zambia. Men were more than twice as likely as women to consume illicit alcohol, while use was higher among older adults, rural residents, and those with limited education. These patterns reflect persistent social and economic inequalities that shape access to safer, regulated beverages. The median ethanol intake among illicit alcohol users reached 62.5 g (IQR: 31.2–156.2 g), revealing not only widespread use but also heavy consumption among certain groups. Together, these findings underscore the scale and complexity of unrecorded alcohol use in Africa, calling for a nuanced discussion on its determinants, health risks, and implications for policy and intervention.

Globally, illicit alcohol use continues to undermine public health systems, especially in low-and middle-income countries where regulation and monitoring are weak ^23^. The World Health Organization estimates that unrecorded alcohol accounts for nearly one-quarter of all alcohol consumed worldwide, exposing millions to preventable harm ^24^. In this study, the prevalence of unrecorded alcohol use across African countries was 8.3%, revealing a significant and ongoing public health challenge. The problem is intensified by weak enforcement of alcohol policies and limited access to affordable, regulated beverages ^25^. Many communities depend on home-brewed or counterfeit drinks that are often more potent and toxic than commercial products. Policymakers should strengthen surveillance systems, invest in laboratory testing, and enforce consumer protection standards. Regional collaboration through the African Union’s Health Strategy can further harmonize regulations and ensure collective efforts to curb illicit production and trade across the continent ^26^.

The study revealed significant differences across African countries, with prevalence ranging from 0.1% in Morocco to nearly 13% in Zambia. Globally, similar disparities exist, often reflecting variations in economic conditions, cultural norms, and enforcement capacity ^16 27^. For instance, regions with strict formal alcohol control but limited enforcement often experience higher levels of unrecorded production and consumption. In Africa, countries with porous borders and informal economies are particularly vulnerable to illicit alcohol circulation ^28 21^. Policymakers should design context-specific strategies that combine regulation with community engagement, such as licensing small-scale brewers or introducing safe production practices. Cross-border coordination through platforms like the East African Community and SADC could help curb illegal trade while promoting safer alternatives that sustain local livelihoods ^29^.

Men were found to be more than twice as likely as women to consume illicit alcohol, aligning with global patterns of higher alcohol use and risk-taking behavior among men ^30 31^. In many African settings, cultural norms reinforce men’s drinking while discouraging women, yet economic hardship and social change are slowly shifting this divide. Older adults and rural populations also show higher rates, possibly due to limited market access to regulated beverages and social acceptance of home-brewed alcohol ^32^. These demographic trends highlight the need for gender- and age-sensitive interventions. Governments should invest in public education, behavior change campaigns, and targeted screening through primary healthcare and community structures ^33^. Empowering women’s groups and youth organizations to promote responsible drinking and safer alternatives could yield long-term public health benefits.

The study’s findings that illicit alcohol use is higher among individuals with lower education and income reflect deep-rooted social inequalities. Globally, poverty and unemployment often push people toward cheaper, unsafe alcohol options, reinforcing cycles of ill health and vulnerability ^34^. Across Africa, weak regulatory systems and limited consumer protection further expose poor communities to harmful products ^35^. Tackling illicit alcohol therefore requires addressing its socioeconomic roots through poverty reduction, job creation, and inclusive health promotion. Policy responses should integrate alcohol control into national social protection programs and community development strategies ^36 37^. Governments could also explore taxation reforms that lower the cost of safe, low-strength beverages to reduce the appeal of dangerous alternatives.

The link between high physical activity and illicit alcohol use likely mirrors the realities of people working in hard manual jobs, where long hours and physical strain push many to rely on cheap, informal alcohol as a quick way to relax or cope^38^. This pattern calls for policies that go beyond promoting healthy lifestyles and instead address the structural conditions shaping alcohol use, such as poor working conditions, limited access to safe recreational spaces, and the easy availability of untaxed alcohol^39^. Practical steps could include workplace wellness programs, targeted screening and brief interventions in high-risk occupations, and stronger enforcement against unsafe informal alcohol markets^40^.

The median ethanol intake of 62.5 g among illicit alcohol users indicates heavy consumption levels that pose serious health risks, from liver damage to injury and addiction. Globally, high-volume unrecorded drinking contributes substantially to the burden of non-communicable diseases and premature deaths ^41^. In Africa, where healthcare access remains limited, such heavy drinking can overwhelm already strained health systems ^42^. Policymakers should prioritize integrating alcohol harm reduction into existing primary healthcare services, including screening, brief interventions, and referral programs. Strengthening community-based monitoring, coupled with clear communication on the dangers of illicit alcohol, can help reduce consumption ^43^. Ultimately, reducing the ethanol content in local brews through safer production practices and quality control could be a pragmatic step toward mitigating the health burden linked to unrecorded alcohol in Africa ^44,44^.

This pattern of higher illicit alcohol use among the self-employed and those not in paid work likely reflects more than the low cost of informal drinks. People working outside formal jobs often rely on loose social networks where informal alcohol is shared, easy to access and part of everyday routines^45,46^. The lack of fixed work hours can also create long periods of unstructured time, making drinking harder to regulate^47^. These dynamics show how economic insecurity, social environments and work patterns interact to shape alcohol use. For policy, this calls for interventions that reach beyond price controls and target the social spaces where informal drinking occurs, such as community-based outreach, support groups and brief interventions tailored to informal workers^48^. Practically, improving livelihood opportunities, strengthening regulation of informal alcohol markets and creating safer community recreation options may help reduce dependence on cheap illicit drinks among these groups^49^.

The beverage-specific results make it clear that the real driver of unrecorded alcohol harm is locally brewed beer and wine. These drinks supply almost all the ethanol consumed, while spirits, imported alcohol, and industrial products contribute very little, even after considering possible underreporting of industrial use. This means the core problem is the informal brewing sector, not counterfeit spirits or smuggled imports. Policy efforts should therefore shift toward safer traditional brewing, simple community-level monitoring, cleaner production practices, and low-barrier licensing or training for local brewers^50^. Targeted health messaging should also focus on the groups most affected, including rural residents, men, those with low education, and people with other high-risk behaviours^51^. By matching interventions to the drinks people actually use, responses become more practical and more likely to reduce harm.

### Strengths and limitations

This study has several important strengths that make its findings useful for both research and policy. It draws on WHO STEPS data collected between 2014 and 2019, which provides nationally representative and comparable information across twelve African countries. This broad coverage gives a reliable picture of illicit alcohol use across different social, economic, and geographic settings. The standardized survey design and robust statistical analysis strengthen the validity of the results and allow for meaningful comparison between groups and countries. The use of multivariable regression and quantile analysis adds depth, highlighting both who is most likely to use illicit alcohol and how heavily they consume it ^52^. These features make the study a valuable evidence base for governments and regional bodies looking to integrate alcohol surveillance into broader non-communicable disease prevention and control strategies.

However, some limitations should be considered when interpreting the findings. Since the study relies on self-reported data, there is a risk of recall and social desirability bias, particularly where alcohol use is socially discouraged or illegal, which may lead to underreporting ^53 54^. The cross-sectional nature of the data limits the ability to draw causal conclusions about the factors driving illicit alcohol use. Furthermore, the analysis used fixed effects for countries to enable direct comparisons, rather than a model with country-specific random effects^55^. While this approach allows for the estimation of individual country-level coefficients, it does not explicitly account for the unobserved heterogeneity between countries, which may influence consumption patterns. This means that some country-specific, unmeasured factors (e.g., cultural norms, enforcement intensity, or informal market dynamics) that affect illicit alcohol use are not modeled as a source of random variance. In addition, the data were collected several years ago, and consumption patterns may have shifted due to recent economic challenges, COVID-19 impacts, or new alcohol policies ^56^. The countries also lacked comprehensive variables, such as biochemical indicators or enforcement data, which limited deeper analysis of health outcomes and regulatory gaps ^57^. Despite these weaknesses, the study highlights the urgent need for stronger national alcohol monitoring systems, regular surveillance, and better coordination across public health, trade, and law enforcement sectors to effectively address the growing problem of illicit alcohol consumption in Africa ^58^.

A notable limitation of this study is that the WHO STEPS survey aggregates illicit alcohol, beer, and wine into a single category of alcoholic beverages^59^. This combined classification limits the ability to assess beverage-specific consumption patterns, associated risk levels, and sociodemographic differences. Important variations in alcohol strength, cost, availability, and cultural preference between these beverage types are therefore not captured^60^. To enhance the precision and policy relevance of future analyses, subsequent rounds of WHO STEPS data collection should consider disaggregating illicit alcohol, beer, wine, and other beverage types, allowing for more detailed monitoring and better-targeted public health interventions.

## Conclusion

In conclusion, this study provides strong evidence that illicit alcohol consumption remains a serious and under-recognized public health challenge across Africa, with clear variations by country, gender, age, and socioeconomic status. The findings underscore the need for governments to move beyond awareness campaigns and adopt comprehensive strategies that combine regulation, enforcement, and social protection. Strengthening national alcohol control policies, improving surveillance systems, and making safer, regulated beverages more accessible are critical steps toward reducing harm. Future efforts should focus on continuous data collection, cross-border collaboration to combat illegal trade, and integration of alcohol harm reduction into broader non-communicable disease programs. By turning this evidence into coordinated action, African countries can protect public health, reduce health inequalities, and move closer to achieving regional and global health targets ^61^.

## Data Availability

The anonymized datasets were accessed and downloaded from the WHO NCD Microdata Repository (https://extranet.who.int/ncdsmicrodata/index.php/catalog/629).

## Abbreviations

## Author contribution

CZ = Conceptualisation, preparing the paper and reviewing it. WFN = Conceptualisation, preparing the paper and reviewing it. Both authors reviewed and approved the final draft.

